# Evaluating and implementing block jackknife resampling Mendelian randomization to mitigate bias induced by overlapping samples

**DOI:** 10.1101/2021.12.03.21267246

**Authors:** Si Fang, Gibran Hemani, Tom G Richardson, Tom R Gaunt, George Davey Smith

**Affiliations:** Population Health Sciences, Bristol Medical School, University of Bristol, Bristol, BS8 2BN, United Kingdom; Medical Research Council (MRC) Integrative Epidemiology Unit (IEU), University of Bristol, BS8 2BN, Bristol, United Kingdom; Novo Nordisk Research Centre, Headington, Oxford, OX3 7FZ, United Kingdom

## Abstract

Participant overlap has been thought to induce overfitting bias into Mendelian randomization (MR) and polygenic risk score (PRS) studies. This hinders the potential research into many unique traits and disease outcomes from large-scale biobanks. Here, we evaluated a block jackknife resampling framework for genome-wide association studies (GWAS) and PRS construction to mitigate the influence of overfitting bias on MR analyses compared to alternative approaches and implemented this study design in causal inference setting using data from the UK Biobank.

We simulated PRS and MR under three scenarios: (1) using weighted SNP estimates from an external GWAS, (2) using weighted SNP estimates from an overlapping GWAS sample and (3) using a block jackknife resampling framework. Based on a conventional *P*-value threshold to derive genetic instruments for MR studies (*P*<5×10^−8^), our block-jackknifing PRS did not suffer from overfitting bias (mean R^2^=0.034) compared to the externally weighted PRS (mean R^2^=0.040). In contrast, genetic instruments derived from overlapping samples explained a higher proportion of variance (mean R^2^=0.048) compared to the externally derived score. The detrimental impact of overfitting bias became considerably larger when using a more liberal *P*-value threshold to construct PRS (e.g., *P*<0.05, mean R^2^=0.103), whereas estimates using jackknife score remained robust to overfitting (mean R^2^=0.084).

In an applied setting, we examined (A) the effects of body mass index on circulating biomarkers and (B) the effect of childhood body size on levels of testosterone in adulthood using methods described above. In the first applied analysis, overlapping sample PRS and block jackknife resampled PRS led to comparable effect sizes, whereas narrower confidence intervals were identified when using the overlapping sample instrument. In the second example, through sex-stratified multivariable and bi-directional MR, we demonstrate that childhood body size indirectly leads to lower testosterone levels in adulthood in males, an effect mediated through adult body size.

**Author summary:** Using genetic variants as instrumental variables for risk factors, Mendelian randomization (MR) provides an approach to explore the genetically predicted effects of modifiable risk factors on disease which is robust to confounding and reverse causation. Genetic instrumental variables are conventionally selected from results of genome-wide association studies on an independent dataset whose sample does not overlap with the dataset being analysed using MR analysis, as this can lead to overfitting bias. This can often be challenging to entirely avoid however, as such association studies are increasingly being performed by meta-analysing several biobanks to achieve the maximum power to detect variants with smaller effect sizes. Moreover, when investigating exposures and outcomes which only a single biobank has measured in sufficiently large samples, avoiding participant overlap requires splitting the study population into subgroups which can limit statistical power. Block jackknife resampling MR provides a solution to conduct causal inference under these circumstances with the maximum statistical power while avoiding bias due to overlapping participants. In this study, we evaluated this study design with simulated dataset in comparison to MR using genetic variants discovered from an external dataset or one with overlapping samples. We applied this approach using UK Biobank to investigate the role of body mass index on circulating biomarkers, as well as the causal relationship between childhood adiposity and testosterone levels in adulthood.

## Introduction

Genome-wide association studies (GWAS) studies have discovered thousands of genetic variants which robustly associate with many different complex traits and disease endpoints in the past two decades. These findings not only lead to potential translatable opportunities for pharmaceutical target development and highlight biological mechanisms and pathways, but also facilitate endeavours in disease prediction and risk stratification using polygenic risk scores (1). Trait-associated genetic variants can also be used as proxies for lifestyle risk factors in Mendelian randomization (MR) studies. Such an approach harnesses genetic data to strengthen causal inference in epidemiological research, and can be implemented in instrumental variables (IV) analyses.(2-4)

MR studies rely on the selection of valid genetic IVs, which are robustly associated with the exposure of interest, affect the outcome only through the exposure being analysed and which do not share a common cause with the outcome (5). Genetic IVs are conventionally selected from an independent dataset whose sample does not overlap with the dataset being analysed using MR analysis, as this can lead to overfitting bias (6). This can often be challenging however, as GWAS are increasingly being performed by meta-analysing several biobanks to achieve the maximum power to detect variants with smaller effects. Furthermore, when investigating exposures and outcomes which only a single biobank has measured in sufficiently large samples, avoiding participant overlap requires splitting the study population into subgroups which can limit statistical power (7). These issues could be avoided using the block jackknife resampling MR, an approach for causal inference in a single sample without participant overlap.

Jackknife resampling, also referred to a leave-one-out or cross-validation, is a form of instrumental variable derivation firstly described by Angrist and colleagues to obtain the fitted value during the first stage of two-stage least squares (2SLS) regression (8). This method can be applied to mitigate the bias in 2SLS conducted in finite-samples when the instruments are weak, namely finite-sample bias or weak instrument bias (6, 8). Different from Angrist et al’s jackknife approach, block jackknife resampling MR applies the jackknife resampling design in blocks to identify genetic instruments robustly associated with the exposure before performing 2SLS to ensure independence between the discovery and applied dataset. Allele scores, or PRS, as a weighted sum of genetic instruments were also constructed in this method to mitigate weak instrument bias in MR using individual level data (9).

In this study, we have evaluated the use of block jackknife resampling to maximize sample sizes for both IV identification and MR analysis using simulated datasets (8). We compared this study design with a one-sample MR design using genetic instruments that were either identified in external GWAS or identified by GWAS on fully overlapping samples. To explore the optimal scenarios for this method, we also carried out a range of simulated MR analyses using allele scores constructed from external GWAS with a range of sample sizes. Next, we evaluated the three methods (IV selection using block jackknife resampling, external GWAS or overlapping GWAS) using real data by analysing body mass index (BMI) data in the UK Biobank (UKB) and the largest BMI GWAS meta-analysis with no reported sample overlap with the UKB cohort. Finally, by applying the block jackknife resampling MR method, we investigated the sex differences in the genetically predicted effect of childhood body size on adult testosterone levels, where both sex-stratified measures were available on the large sample available in UKB. The influence of childhood body size on adult testosterone levels was then investigated in terms of direct and indirect effects after accounting for adult body size, by extending the jackknifing study design into a multivariable setting.

## Description of method

Block jackknife resampling MR is a modified approach for one-sample MR which uses individual-level genotype and phenotype data to study the causal relationship between risk factors (exposures) and diseases or traits (outcomes) in a single dataset. It provides maximum statistical power for causal inference while avoiding biases due to participant overlap between the datasets for IV discovery and for causal inference.

Performing a block jackknife resampling MR study involves the below steps:

1. Split the full dataset randomly into N groups.
2. Perform GWAS on the exposure of interest using all samples from each possible set of N-1 groups to obtain SNPs (*P*<5×10^−8^) and weights for PRS construction for each set.
3. Construct PRS for the exposure for individuals in the remaining group for each set, so that PRS is constructed for every individual in the full dataset.
4. Combine all groups and use PRS as genetic IV for the exposure in one-sample MR using 2SLS regressions.

With the availability of additional phenotype data, the block jackknife resampling framework could be extended to identify direct and indirect effects from the exposure of interest in a multivariable setting. This approach could also be easily applied to detect reverse causality when GWAS is performed for and PRS constructed on the outcome.

## Verification and Comparison

### Simulated analysis 1: Bias brought by overlapping samples in simulated data

To investigate the effect of sample overlap and the advantages of using a block jackknife resampling framework to mitigate overfitting bias, we performed extensive simulations to construct PRS under the combination of three GWAS frameworks and 13 different p-value thresholds. **Figure 1** shows a schematic illustration of the three approaches.

**Figure 1:**
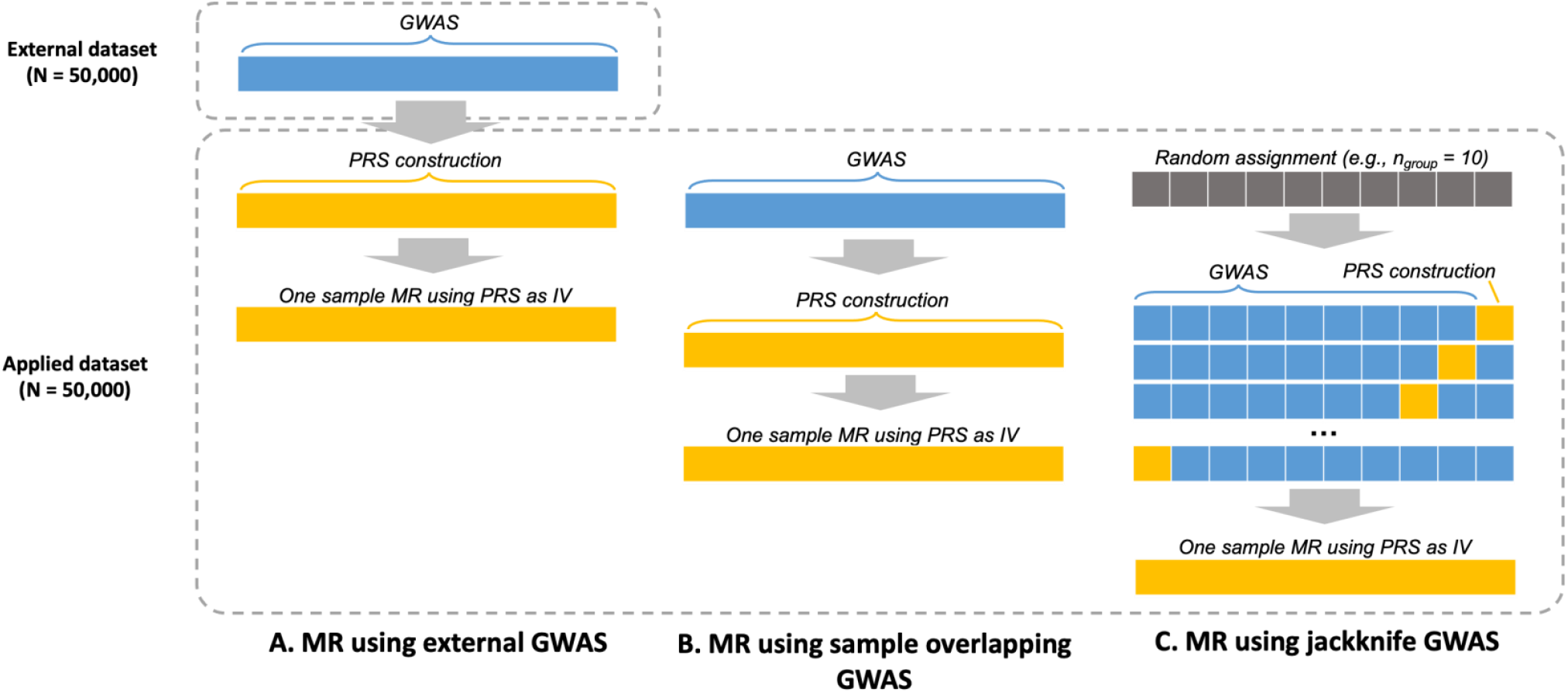
Schematic diagram showing the three frameworks for selecting samples for conducting genome-wide association studies (GWAS) and polygenic risk scores (PRS) construction in the simulated analyses. The three frameworks applied in the simulation analyses are (A) Mendelian Randomization (MR) using external GWAS: SNPs and their weights used for PRS construction was identified from a GWAS on an external dataset, different from the applied dataset which is used for PRS construction and MR; (B) MR using sample overlapping GWAS: the same dataset is used for effect estimate, PRS construction and MR; (C) MR using jackknife GWAS: effect estimate, PRS construction and MR under the block jackknife resampling framework. Blue: dataset used to estimate SNP effects; Yellow: dataset used to construct PRS and carry out one-sample MR.

We generated 1000 pairs of simulated datasets consisting of n=50000 (referred to as our ‘applied dataset’) and n=50000 (referred to as our ‘external dataset’) individuals. In the applied datasets, three continuous phenotypes (i.e., our exposure, outcome, and confounder) and genetic data consisting of 500 independent SNPs were simulated for every individual. In the external datasets, two continuous phenotypes (i.e., our exposure and confounder) and genetic data consisting of 500 SNPs were simulated for every individual. Data simulation was conducted using the “simulateGP” R package (https://github.com/explodecomputer/simulateGP/). A set of fixed parameters were applied, including the total variance in the exposure explained by all SNPs (*R*^2^ = 0.1), minor allele frequency (MAF) of all SNPs (0.2), the effect from the exposure to the outcome (*β*_*XY*_ = 0.2), the effect from the covariate to the exposure (*β*_*UX*_ = 0.4) and the effect from the covariate to the outcome (*β*_*UY*_ = 0.3).

Using the simulated data described above, GWAS of the exposure variables were conducted (A) using the external dataset (referred to as ‘external GWAS’), (B) using all samples in the applied dataset (referred to as ‘overfitted GWAS’), and (C) following a block jackknife resampling framework in the applied dataset (referred to as ‘jackknife GWAS’) (**Figure 1**). GWAS on simulated data were performed using linear regression as implemented through the “gwas” function in the “simulateGP” R package.

For individuals in the applied datasets, three PRSs were constructed as the weighted sum of effect alleles that reached the *P*-value threshold in the GWAS summary statistics generated by the three approaches. Each of the 13 scenarios corresponded to a *P*-value threshold of 5×10^−8,^ 1×10^−7,^ 1×10^−6^, 1×10^−5^, 1×10^−4^, 5×10^−4^, 1×10^−3^, 5×10^−3^, 0.01, 0.05, 0.1, and 1, respectively. LD clumping was not undertaken on simulated GWAS statistics because all SNPs were simulated to be uncorrelated.

The adjusted correlation coefficient 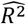 between PRS and the exposure were calculated using linear regression in the applied dataset for each of the 3 simulated PRS in turn. Estimates (beta coefficients 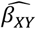, and their standard errors 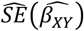) of the genetically predicted effect of exposures on the simulated outcomes were calculated via two-stage least squares (2SLS) regression. This was achieved using the ivreg() function from the ‘ivpack’ R package. The R package ‘rsimsum’ (10) was used to compute simulation metrics include the mean, median, bias, empirical standard error, percentage gain in precision relative to the externally weighted PRS, mean squared error for adjusted 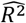 and 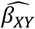, the mean, median and relative percentage error in standard error for 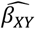, the model-based standard error and relative error in model-based standard error for 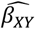, the coverage and bias-eliminated coverage for, and Monte Carlo standard error of all summary statistics.

An overview of the performance of simulated PRS is presented in **Figure 2**. Adjusted correlation coefficient 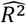 between PRS and the exposure phenotype represents the predictive ability of the PRS (**Figure 2A**). Using a conventional p-value threshold to derive genetic instruments for MR studies (i.e., *P*<5×10^−8^), our block jackknife resampling PRS did not appear to suffer from overfitting bias (mean 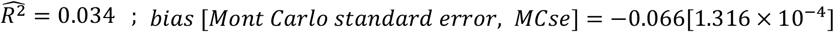) in comparison to the externally weighted PRS (mean 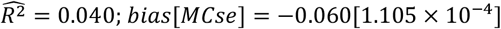). However, genetic instruments derived from overlapping samples typically explained a higher proportion of variance (mean 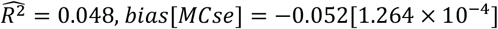) compared to the externally derived score. The detrimental impact of overfitting bias became considerably larger when using a more liberal *p*-value threshold to construct PRS (e.g., when *P* = 0.05, mean 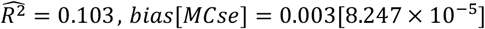), whereas block jackknife resampled estimates remained robust to overfitting (when *p* = 0.05, mean 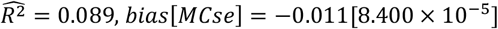).

**Figure 2.**
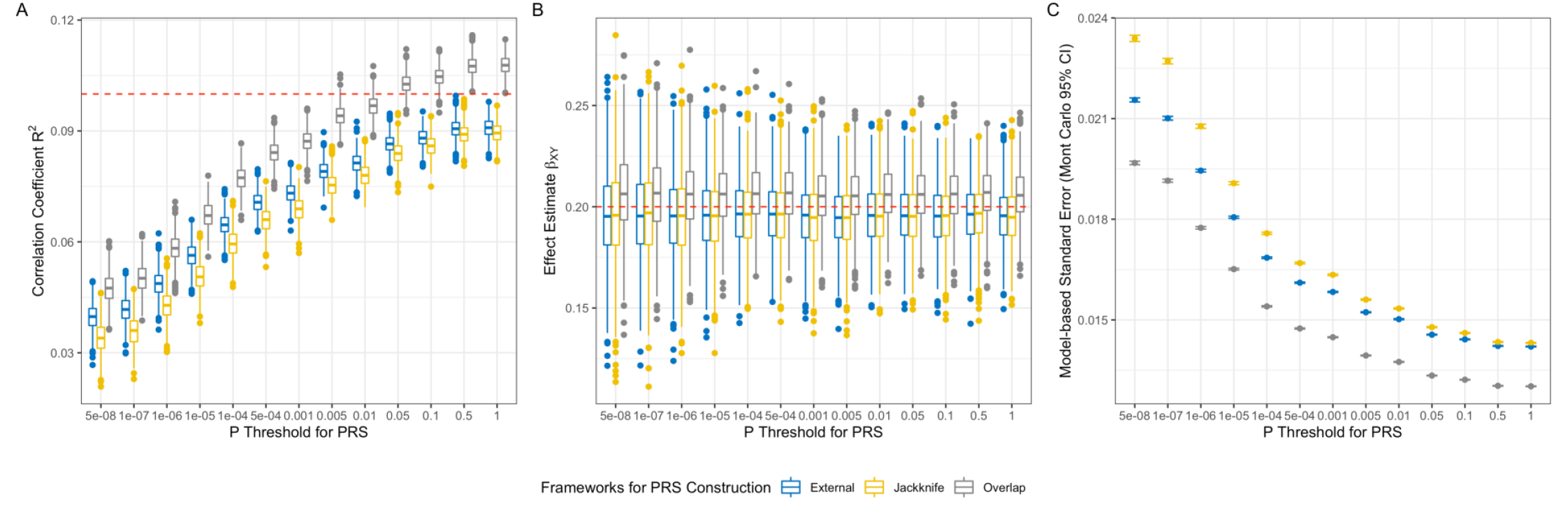
Results from simulation studies comparing three methods for GWAS and PRS construction for prediction and Mendelian randomization (MR). The performance of PRS constructed under three GWAS frameworks in phenotype prediction (A) and one-sample Mendelian randomization (B and C). (A) Boxplots showing the distribution of the correlation between simulated PRS and exposure. (B) Boxplots showing the distribution of the estimates (beta coefficients) of effect from the exposure to the outcome. (C) Scatter plot showing the model-based standard errors of the effect estimates in plot (B) and their Mont Carlo 95% confidence intervals (95% CI). Red lines represent the parameters used in data simulation, i.e., the true value of correlation coefficient R^2^ (0.10) in plot A and beta coefficient (0.20) in plot B.

In one-sample MR, exposures instrumented by PRS constructed with overlapping samples showed slightly inflated effects on the outcome under all p-value thresholds (overall mean 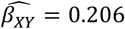, overall *bias* = 0.006), whereas block jackknife resampled PRS and externally weighted PRS both suggested effects slightly biased towards the null (both overall mean 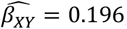; overall *bias* = −0.004) (**Figure 2B**). Moreover, results derived using overlapping sample PRS had lower model-based standard errors for the effect estimate, whereas block jackknife resampled PRS produced results with higher standard error compared with externally weighted scores (**Figure 2C**).

For the full summary statistics of this simulated analysis see **Supplementary Table S1 and S2**.

### Simulated analysis 2: Differing sample sizes for external GWAS

Many published trait and disease GWAS do not include UKB, but these typically have smaller sample sizes, which is a key determinant of the power of GWAS and the number of trait-associated SNPs detectable from such studies (11). Thus, to further evaluate the optimal situation for applying the block jackknife resampling MR, we performed another simulation analysis to compare it with classic one-sample MR using PRS constructed with SNPs identified through external GWAS with a small sample size (compared to the applied dataset) as the genetic instruments for the exposure.

Using the same approach used in the first simulated analysis, we generated phenotype and genotype data for 1000 pairs of applied datasets (n=50000) and external datasets with each of the different sample sizes (n=10000, 15000, 20000, 25000, 30000, 35000, 40000, 45000 and 50000). GWAS and PRS construction were undertaken using the three frameworks performed in the first analysis, except all PRS were calculated using a stringent *P*-value threshold for MR (*P*<5×10^−8^). Linear regression and 2SLS were performed to generate the correlation coefficients and estimate of the effects from the exposure to the outcome. The R package ‘rsimsum’ (10) was used to compute simulation metrics mentioned in the first analysis.

In general, the adjusted correlation coefficient 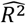 between PRS and exposure increased with the increase of the sample size of external GWAS in results generated using externally weighted PRS. Using a MR level *P*-value threshold (i.e., *P*<5×10^−8^), block jackknife resampling PRS on an applied dataset of 50,000 individuals explained higher variance in the exposure (mean 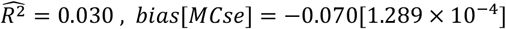) compared with a weighted score generated using external GWAS with a sample size of 40,000 or less (e.g., when n=40,000, mean 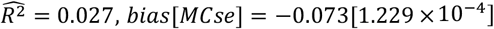).

In one-sample MR, median effect estimates *β*_*XY*_ are relatively consistent across results generated with different external GWAS size and the block jackknife resampling framework. However, exposures instrumented by PRS constructed with the three GWAS with the smallest sample size (n=10,000, 15,000 or 20,000) exhibited high variations in the estimate of effects from the exposure to the outcome (e.g., when n=10,000, mean 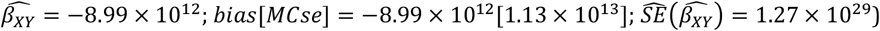).

For the full simulation metrics see **Supplementary Table S3 and S4**.

## Applied examples

### Applied example 1: Effect of body mass index on circulating biomarkers

In the first applied analysis, we implemented the block jackknife resampling MR together with the two other approaches used in the simulated analysis to examine the causal effects from body mass index on a set of 12 circulating biomarkers using data from the UK Biobank (UKB).

#### Methods

UKB is a large-scale prospective cohort study consisting of approximately 500,000 individuals aged between 38 and 73 years at baseline from across the United Kingdom (12). Data were collected based on clinical examinations, assays of biological samples, questionnaires and interviews, as well as genome-wide genotyping as described previously (13, 14). UK Biobank received ethical approval from the Research Ethics Committee (REC reference 11/NW/0382).

Body mass index (BMI) was calculated by weight (kg) divided by standing height (m) squared, both measured at the initial assessment. We focused on a subset of circulating biomarkers which were all measured from serum samples also obtained at baseline. (C-reactive protein, alkaline phosphate, testosterone, glucose, cystatin C, urea, urate, albumin, direct bilirubin, total bilirubin, gamma glutamyltransferase, alanine aminotransferase, aspartate aminotransferase).

In this applied analysis, we constructed a BMI PRS using data from UKB participants (n=333,894) with BMI associated genetic variants identified and their weights estimated using three GWAS frameworks (**Figure 1**):

1. Using summary statistics of the Locke et al. BMI GWAS meta-analysis (15) which has no reported sample overlap with the UKB to generate an externally weighted PRS;
2. Using the full UKB cohort to perform BMI GWAS and generate sample overlapping PRS;
3. Using the full UKB cohort to perform BMI GWAS with the block jackknife resampling framework and generating a block jackknife resampled PRS.

In scenario 1, the weighted PRS for BMI was constructed using genome-wide significant (*P*<5×10^−8^) genetic variants from the summary statistics of the Locke *et al*. BMI GWAS meta-analysis on up to 322,154 individuals of European descent. In scenario 2, a GWAS for BMI was conducted on 461,377 UKB participants using a linear mixed model (LMM) association method as implemented in BOLT-LMM (v2.3) (16) to account for population structure in the UKB. Age, sex and genotyping chip were included as covariates. The GWAS results were clumped using a reference panel consisting of a subset of unrelated UK Biobank participants of European ancestry (N=10,000), as described in a previous study (17) to identify independent genetic variants (linkage disequilibrium threshold r^2^<0.001 within a 1000kb region) which reached the genome-wide significance (*P*<5×10^−8^). Those independent variants were used to construct weighted PRS for BMI. Clumping and PRS construction were achieved using PLINK (v2.0) (18). In scenario 3, we first randomly assigned the 461,377 UKB participants with BMI phenotype into 10 groups. GWAS was undertaken on each possible set of nine groups with adjustment for the same covariates as in scenario 2. GWAS results were clumped and then used to generate weighted PRS for individuals in the remaining group for each set. Details of the GWAS pipeline can be found in **Supplementary Methods**.

Using a BMI PRS generated through three frameworks as described above, we performed one-sample MR to examine the relationship between BMI and 12 circulating biomarkers (C-reactive protein, alkaline phosphate, glucose, cystatin C, urea, urate, albumin, direct bilirubin, total bilirubin, gamma glutamyltransferase, alanine aminotransferase, aspartate aminotransferase) in the UKB. Biomarker levels underwent rank-based inverse normal transformation before MR analysis to ensure their normality. We applied 2SLS regressions using the BMI PRS as a genetic instrument, with sex, age and the first 10 principal components fitted as covariates. This allowed us to compare estimates from the 3 different PRS generated using each of the scenarios described above.

#### Results

One-sample MR using three sets of BMI PRS provided generally consistent evidence in the effect from BMI on circulating biomarkers of interest, as shown in forest plots in **Figure 3**.

**Figure 3.**
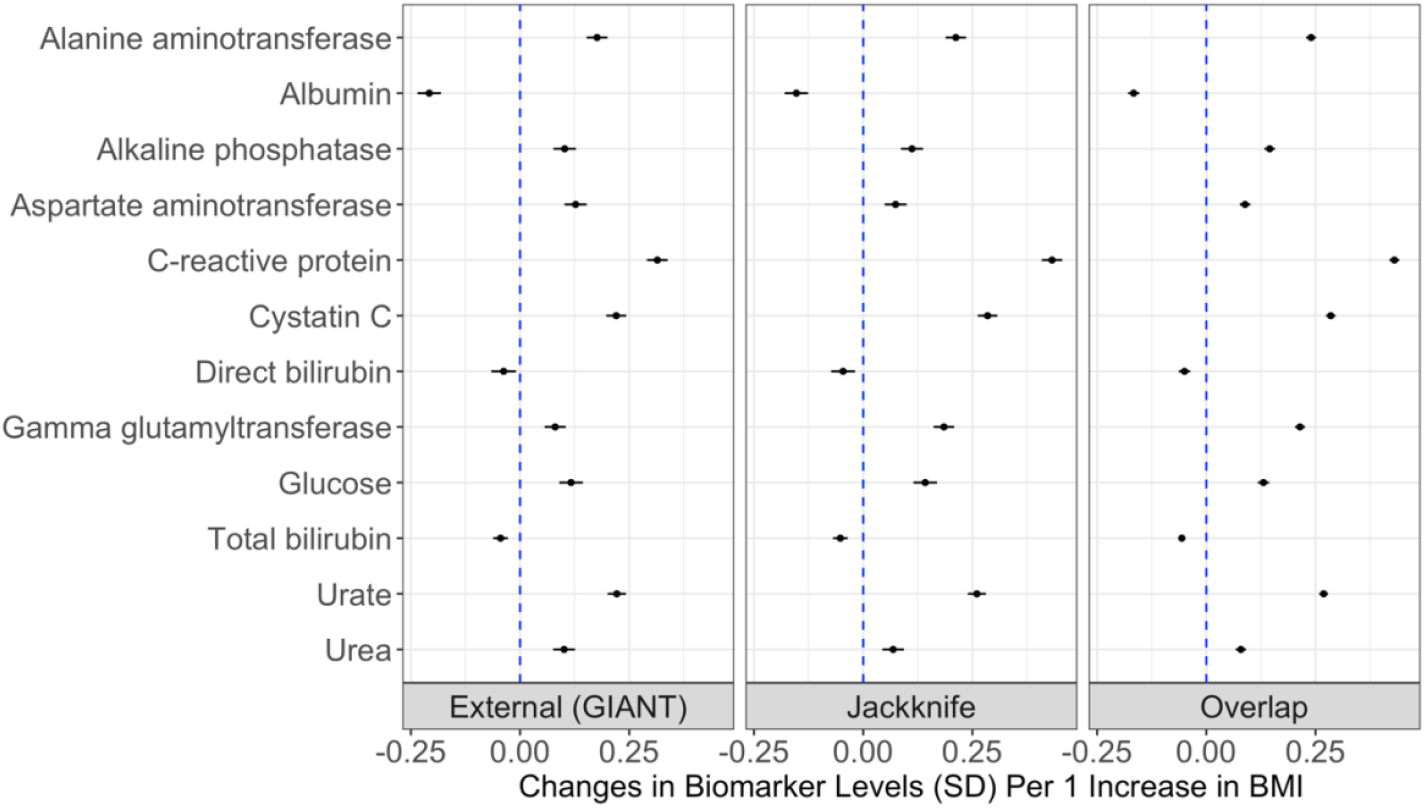
Univariable one-sample MR estimates from body mass index (BMI) and 12 serum biomarkers. One-sample MR estimates and their 95% confidence intervals of effects from the genetic liability towards a high BMI on the levels of 12 biomarkers in the UK Biobank study. External (GIANT) refers to the results generated using externally weighted BMI PRS constructed with Locke et al. GWAS summary statistics. Jackknife refers to the results generated using block jackknife resampling BMI PRS constructed with UK Biobank (UKB) data. Overlap refers to the results generated using internally weighted BMI PRS constructed with sample overlapping GWAS on UKB participants.

BMI provided evidence of a genetically predicted effect on all 12 biomarkers (based on false discovery rates (FDR) < 5%), with C-reactive protein (CRP) having the strongest evidence of an effect in all scenarios (externally weighted PRS: Beta=0.31 SD change in the levels of CRP per 1 increase in BMI PRS, 95% confidence interval (CI)=0.29 to 0.34, p=4.57×10^−147^; block jackknife resampled PRS: Beta=0.43, 95% CI=0.41 to 0.46, p=1.96×10^−288^; sample overlapping PRS: Beta=0.43, 95% CI=0.42 to 0.44, p<1×10^−300^). The effect estimates for BMI on each of the 12 biomarkers were generally consistent between three PRSs (**Supplementary Table S5**). In contrast, effect estimates for BMI instrumented using overlapping the sample score typically had much smaller standard errors (average 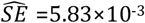), resulting in narrower confidence intervals compared with the other two sets of PRS (externally weighted PRS: average 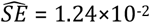; block jackknife resampled PRS: average 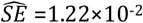).

PRS generated by the three methods all provided strong instruments for BMI. Their *F*-statistics ranged between 4707 and 5695 when using PRS constructed by the Locke. et al GWAS, between 5000 and 5857 when using block jackknife resampled PRS and between 23339 and 27336 when using PRS generated by GWAS on overlapping samples.

### Applied example 2: Effect of childhood adiposity on adult testosterone levels

We applied block jackknife resampling MR to investigate sex-specific effects of childhood adiposity on serum testosterone levels measured in adults in the UK Biobank through (1) univariable MR, (2) multivariable MR, and (3) bi-directional MR, all in a one-sample setting.

#### Methods

Childhood body size was derived using questionnaire data asking participants to recall their body size at 10 years old as ‘thinner’ or ‘plumper’ than average, or ‘about average’, as described previously (19). For comparative purposes, adult body size was derived by categorizing the BMI data into a 3-category variable using the same proportions as seen in the strata of the childhood body size variable. Before analysis, the UKB measurement of circulating testosterone was stratified by sex and then transformed using a rank-based inverse normal transformation.

Block jackknife resampling was conducted in female-only and male-only UKB participants separately to construct sex-specific PRS for childhood and adult body size as well as the levels of circulating testosterone. UKB samples were randomly assigned into 10 groups for both males and females. GWAS on the three phenotypes were undertaken on participants from each of the nine groups adjusted for age and genotyping chip, and the results were used to construct PRS for individuals in the remaining group. The robustness of using allele scores constructed with SNPs associated with this childhood body size phenotype as the instrument for childhood adiposity has been validated in three independent cohorts in previous analyses (19-21).

Next, we performed univariable one-sample MR using sex-specific PRS for childhood body size as genetic instruments. 2SLS regressions were undertaken for males and females separately, where age and the first 10 principal components were fitted as covariates. To assist with comparisons between adult and childhood body size, we also estimated the effects from the comparable 3-tier adult body size variable on testosterone levels using the univariable model.

Moreover, we used multivariable one-sample MR to estimate the direct and indirect role of childhood body size on testosterone levels where evidence of a total effect was identified in univariable analyses. This was achieved by accounting for adult body size as an additional exposure in the 2SLS analysis.

Finally, we performed one-sample MR in the reverse direction, i.e., using the levels of testosterone as the exposure and childhood/adult body size as the outcome, to examine whether genetically predicted levels of testosterone in adulthood influences either childhood body size (i.e. as a negative control) or adult body size. One-sample MR using 2SLS were performed, where age and the first 10 principal components were fitted as covariates. Results were obtained from MR using block jackknife resampling PRS and using overlapping sample PRS (constructed using SNPs and weights from a GWAS on all UKB participants) as the genetic instruments for the levels of circulating testosterone.

All statistical analysis were undertaken using R (v4.0.2) (22)

#### Results

Univariable one sample MR provided strong evidence of a genetically predicted effect between a childhood body size and the levels of testosterone in males (Beta=-0.40 SD change in the levels of testosterone per 1 increase in body size category, 95% CI=-0.51 to −0.29, p=4.56×10^−14^), whereas there was little evidence of an effect in females (Beta=3.19×10^−3^, 95% CI=-0.09 to 0.09, p=0.945) (**Figure 4A)**. We also found evidence of an effect of genetically predicted adult body size on the levels of testosterone in both sexes based on univariate MR. Higher adult body size had a genetically predicted effect on lower testosterone levels in males (Beta=-0.48, 95% CI=-0.56 to-0.39, p=4.01×10^−29^), whereas the opposite direction of effect on testosterone was found in females (Beta=0.016, 95% CI=0.05 to 0.27, p=0.006) (**Figure 4A)**.

**Figure 4.**
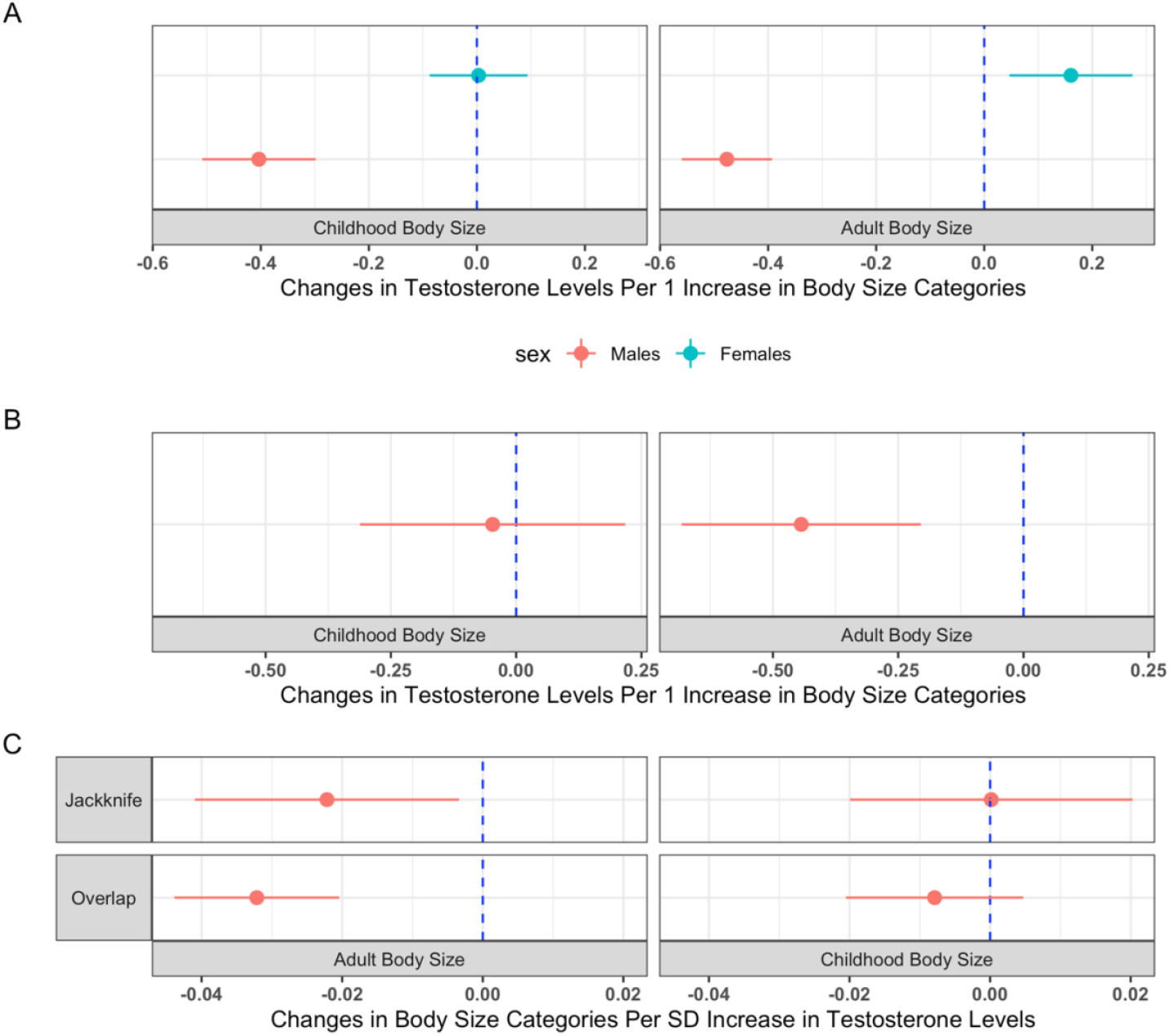
Univariable and multivariable MR on the direct and indirect effects from childhood body size on circulating testosterone levels. (A) Sex-stratified univariable one-sample MR estimates between childhood and adult body sizes on circulating testosterone levels in males and females. (B) Multivariable one-sample MR estimates between childhood and adult body sizes on circulating testosterone levels in males. (C) One-sample MR estimates between the levels of testosterone on childhood and adult body sizes in males, using block jackknife resampling PRS (“Jackknife”) and sample overlapping PRS (“Overlap”) for the levels of testosterone as genetic instruments. SD, standard deviation.

The testosterone lowering effect of higher genetically predicted childhood body size observed in males in the univariable analysis was further explored in a multivariable setting. Results from multivariable MR did not support a direct effect of childhood body size on adulthood testosterone levels after accounting for the genetically predicted effect of adult body size in males (Beta=-0.05 per 1 increase in childhood body size category when accounting for adult body size, 95% CI=-0.31 to 0.22, p=0.728). However, there was evidence for an indirect negative effect putatively mediated along the causal pathway involving adult body size (Beta=-0.44 per 1 increase in adult body size category when accounting for childhood body size, 95% CI=-0.68 to −0.20, p=2.66×10^−4^) (**Figure 4B)**.

The relationship between body size and the levels of testosterone in males was further assessed using MR in the reverse direction. Results from MR using block jackknife resampling PRS as the genetic instrumental variable provided little evidence for an effect from genetically predicted higher levels of testosterone on childhood body size (Beta=1.62×10^−4^ per 1 SD increase in the levels of testosterone, 95% CI=-0.02 to 0.02, p=0.987) whilst they supported a marginal effect on adult body size (Beta=-0.02, 95% CI=-0.04 to −3.38×10^−3^, p=0.021). Despite the slight differences in effect estimates, MR using PRS constructed using GWAS on overlapping samples provided results consistent with those from our method (**Supplementary Table S6C**) (**Figure 4C)**.

Block jackknife resampling PRS provided strong instruments for childhood and adult body size as well as the levels of testosterone. In females, the F-statistics for childhood and adult body size in the univariable setting are 1003 and 581 respectively. In males, the conditioned F-statistics (in multivariable MR) are 629 and 872 for childhood and adult body size respectively. In reverse MR analysis, the F-statistics for the levels of testosterone in males is 4342. PRS generated using GWAS on overlapping samples provided a F-statistics of 11313 for the levels of testosterone in males. Detailed results from the applied example 2 are in **Supplementary Table S6**.

## Discussion

Sample overlap between multiple GWAS studies is becoming increasingly prevalent due to the growth of GWAS meta-analyses in recent years. When estimating the causal effect of exposures on continuous outcomes using MR approaches such as 2SLS or the inverse-variance weighted methods, bias due to sample overlap was shown to be linearly correlated with the percentage of overlap between samples in previous literature (6). However, this bias was not found for binary outcomes when the first-stage regression of 2SLS is conducted using risk factor data on individuals in the control group (6). In this study, we examined biases due to participant overlap in one-sample MR results on a continuous outcome and evaluated the effectiveness of a block jackknife resampling one-sample MR method in mitigating this bias with both simulated datasets and real data in the UKB. By applying block jackknife resampling MR to investigate the causal effect of genetically predicted childhood body size on adult serum testosterone levels, we demonstrated the application of this approach in terms of enabling sex-stratified, multivariable and bi-directional MR analyses in the absence of an external dataset.

Having an elevated BMI has been linked to multiple health conditions and disease endpoints in multiple observational and MR studies (23-28). Using the block jackknife resampling MR approach outlined in this manuscript, we were able to replicate previous identified effects of higher BMI on the levels of various serum biomarkers which are routinely measured in a clinical setting, including C-reactive protein (29), cystatin C (30), alanine aminotransferase and gamma glutamyltransferase (31) as well as urate (32). Together with results from simulation analyses, this first applied example on BMI and biomarkers validated the robustness of block jackknife resampling MR in providing equivalent causal inference compared with classic MR using individual level data and externally identified genetic instruments. Our findings from simulated data also suggest that the jackknife resampling study design is preferable in situations where (a) you have a large dataset containing both exposure and outcome data that can be used to derive genetic instrument variables and perform MR and (b) the only available external GWAS is substantially smaller.

Despite the increase in GWAS meta-analysis for common traits and diseases such as BMI and type 2 diabetes, numerous phenotypes are under-investigated in GWAS and MR due to limited data available in non-overlapping samples. In the second applied example undertaken by our study, we examined the sex-specific causal effect from childhood adiposity on adult serum testosterone levels, as an illustration of how block jackknife resampling MR could be applied to study causal relationships involving phenotypes with limited data sources. Testosterone is a sex hormone produced predominately in males and plays important roles in the development of masculine characteristics. Low levels of testosterone produced in the body, namely testosterone deficiency or hypogonadism, is a condition that primarily affects older men (33). This condition is often treated with testosterone replacement therapy, however, concerns regarding the safety of this therapy have been raised as adverse events following its use have been reported (34-36). Therefore, identifying any modifiable risk factors contributing to changes in testosterone levels would be valuable for preventing this condition. Childhood-onset adiposity, previously identified as an early life risk factor for multiple cardiovascular diseases and cancer (37, 38), was reported to associate with lower testosterone levels in adulthood in an observational study (39). This association could be due to confounding and thus should be further studied using MR to determine whether childhood adiposity has a causal effect on lower levels of testosterone.

Independent SNPs that reached genome-wide significance (*P*<5×10^−8^) in GWAS are often selected as candidate IVs for MR. GWAS with a larger sample size usually have more power to identify SNPs associated with traits or diseases, providing stronger instruments for MR. Meanwhile, a dataset for one-sample MR analysis also needs to be large enough to provide statistical power for identifying any causal relationship between the exposure and the outcome. To date, the largest datasets on childhood adiposity (proxied by recalled body size compared to others at 10 years old) and serum levels of testosterone were both available in UKB. With the advantage of a large sample size, GWAS on UKB traits can provide higher statistical power for subsequent MR analysis by enabling use of a larger number of instruments compared to those available externally, but they have the issue of participant overlap between the samples for IV discovery and MR. Instead, using block jackknife resampling MR, we were able to identify sex-specific genetic IVs for childhood body size and examine the sex-stratified causal effect from childhood adiposity on adult serum testosterone using the largest dataset available without violating the independence assumption of MR or losing much power. Univariable and multivariable MR support an indirect causal effect of higher childhood body size on lower testosterone in men mediated by higher adult body size, consistent with a previous finding that higher adult BMI is causally associated with lower testosterone levels in men (40). This suggests that the influence of childhood obesity on lower serum testosterone could be mitigated if one loses weight in adulthood, similar to previously identified causal effects of early life and adult body size on type 2 diabetes and risk of coronary artery disease (19). This applied example illustrated the value of this block jackknife resampling study design in a causal inference setting when using limited sources of data for sex stratified IV discovery, as well as when applying MR in multivariable and bi-directional settings.

This method has some limitations. Firstly, to identify block jackknife resampled genetic instruments requires higher levels of computational power than in classic MR where IVs were identified in one GWAS study. Secondly, through block jackknife resampling MR we cannot directly identify the existence of horizontal pleiotropic SNPs. Horizontal pleiotropy arises when one or more SNPs used as the genetic instrumental variable for the exposure influence the outcome through a pathway which does not involve the exposure. The inclusion of such SNPs as genetic instrumental variables for an exposure can reintroduce confounding and lead to bias in causal inference (5). Horizontal pleiotropy could be detected through detailed examination of individual SNPs used for PRS construction in each block separately, through evaluating the between-SNP heterogeneity in the ratio of associations between genotype and the outcome and the exposure (41, 42).

In summary, block jackknife resampling MR method provides researchers with an approach to perform hypothesis testing with limited sources of data before conducting a comprehensive assessment of causal relationships between modifiable risk factors and complex disease traits and outcomes.

## Data Availability

All data produced in the present work are contained in the manuscript.

## Acknowledgement

Quality Control filtering of the UK Biobank data was conducted by R. Mitchell, G. Hemani, T. Dudding, L. Corbin, S. Harrison, L. Paternoster as described in the published protocol (43). The MRC IEU UK Biobank GWAS pipeline was developed by B. Elsworth, R. Mitchell, C. Raistrick, L. Paternoster, G. Hemani, T. Gaunt (44). We would also like to thank the participants of the UK Biobank study.

## References

1. Tam V, Patel N, Turcotte M, Bosse Y, Pare G, Meyre D. Benefits and limitations of genome-wide association studies. Nat Rev Genet. 2019;20(8):467–84.

2. Smith GD, Ebrahim S. ‘Mendelian randomization’: can genetic epidemiology contribute to understanding environmental determinants of disease? Int J Epidemiol. 2003;32(1):1–22.

3. Lawlor DA, Harbord RM, Sterne JA, Timpson N, Davey Smith G. Mendelian randomization: using genes as instruments for making causal inferences in epidemiology. Stat Med. 2008;27(8):1133–63.

4. Richmond RC, Davey Smith G. Mendelian Randomization: Concepts and Scope. Cold Spring Harb Perspect Med. 2021.

5. Davey Smith G, Hemani G. Mendelian randomization: genetic anchors for causal inference in epidemiological studies. Hum Mol Genet. 2014;23(R1):R89-98.

6. Burgess S, Davies NM, Thompson SG. Bias due to participant overlap in two-sample Mendelian randomization. Genet Epidemiol. 2016;40(7):597–608.

7. Sadreev II, Elsworth BL, Mitchell RE, Paternoster L, Sanderson E, Davies NM, et al. Navigating sample overlap, winner’s curse and weak instrument bias in Mendelian randomization studies using the UK Biobank. medRxiv. 2021:2021.06.28.21259622.

8. Angrist JD, Imbens GW, Krueger AB. Jackknife instrumental variables estimation. Journal of Applied Econometrics. 1999;14(1):57–67.

9. Burgess S, Thompson SG. Use of allele scores as instrumental variables for Mendelian randomization. Int J Epidemiol. 2013;42(4):1134–44.

10. Koehler E, Brown E, Haneuse SJ. On the Assessment of Monte Carlo Error in Simulation-Based Statistical Analyses. Am Stat. 2009;63(2):155–62.

11. Spencer CC, Su Z, Donnelly P, Marchini J. Designing genome-wide association studies: sample size, power, imputation, and the choice of genotyping chip. PLoS Genet. 2009;5(5):e1000477.

12. Sudlow C, Gallacher J, Allen N, Beral V, Burton P, Danesh J, et al. UK biobank: an open access resource for identifying the causes of a wide range of complex diseases of middle and old age. PLoS Med. 2015;12(3):e1001779.

13. Collins R. What makes UK Biobank special? The Lancet. 2012;379(9822):1173–4.

14. Bycroft C, Freeman C, Petkova D, Band G, Elliott LT, Sharp K, et al. The UK Biobank resource with deep phenotyping and genomic data. Nature. 2018;562(7726):203–9.

15. Locke AE, Kahali B, Berndt SI, Justice AE, Pers TH, Day FR, et al. Genetic studies of body mass index yield new insights for obesity biology. Nature. 2015;518(7538):197–206.

16. Loh PR, Tucker G, Bulik-Sullivan BK, Vilhjalmsson BJ, Finucane HK, Salem RM, et al. Efficient Bayesian mixed-model analysis increases association power in large cohorts. Nat Genet. 2015;47(3):284–90.

17. Kibinge NK, Relton CL, Gaunt TR, Richardson TG. Characterizing the Causal Pathway for Genetic Variants Associated with Neurological Phenotypes Using Human Brain-Derived Proteome Data. Am J Hum Genet. 2020;106(6):885–92.

18. Chang CC, Chow CC, Tellier LC, Vattikuti S, Purcell SM, Lee JJ. Second-generation PLINK: rising to the challenge of larger and richer datasets. GigaScience. 2015;4(1).

19. Richardson TG, Sanderson E, Elsworth B, Tilling K, Davey Smith G. Use of genetic variation to separate the effects of early and later life adiposity on disease risk: mendelian randomisation study. BMJ. 2020;369:m1203.

20. Richardson TG, Mykkanen J, Pahkala K, Ala-Korpela M, Bell JA, Taylor K, et al. Evaluating the direct effects of childhood adiposity on adult systemic metabolism: a multivariable Mendelian randomization analysis. Int J Epidemiol. 2021.

21. Brandkvist M, Bjorngaard JH, Odegard RA, Asvold BO, Smith GD, Brumpton B, et al. Separating the genetics of childhood and adult obesity: a validation study of genetic scores for body mass index in adolescence and adulthood in the HUNT Study. Hum Mol Genet. 2021;29(24):3966–73.

22. R Core Team. R: A Language and Environment for Statistical Computing. Vienna, Austria: R Foundation for Statistical Computing; 2020.

23. Gomez-Ambrosi J, Silva C, Galofre JC, Escalada J, Santos S, Gil MJ, et al. Body adiposity and type 2 diabetes: increased risk with a high body fat percentage even having a normal BMI. Obesity (Silver Spring). 2011;19(7):1439–44.

24. Ortega FB, Lavie CJ, Blair SN. Obesity and Cardiovascular Disease. Circ Res. 2016;118(11):1752–70.

25. Avgerinos KI, Spyrou N, Mantzoros CS, Dalamaga M. Obesity and cancer risk: Emerging biological mechanisms and perspectives. Metabolism. 2019;92:121–35.

26. Larsson SC, Back M, Rees JMB, Mason AM, Burgess S. Body mass index and body composition in relation to 14 cardiovascular conditions in UK Biobank: a Mendelian randomization study. Eur Heart J. 2020;41(2):221–6.

27. Bull CJ, Bell JA, Murphy N, Sanderson E, Davey Smith G, Timpson NJ, et al. Adiposity, metabolites, and colorectal cancer risk: Mendelian randomization study. BMC Med. 2020;18(1):396.

28. Carreras-Torres R, Johansson M, Gaborieau V, Haycock PC, Wade KH, Relton CL, et al. The Role of Obesity, Type 2 Diabetes, and Metabolic Factors in Pancreatic Cancer: A Mendelian Randomization Study. J Natl Cancer Inst. 2017;109(9).

29. Timpson NJ, Nordestgaard BG, Harbord RM, Zacho J, Frayling TM, Tybjaerg-Hansen A, et al. C-reactive protein levels and body mass index: elucidating direction of causation through reciprocal Mendelian randomization. Int J Obes (Lond). 2011;35(2):300–8.

30. Xu X, Eales JM, Jiang X, Sanderson E, Scannali D, Morris AP, et al. Obesity as a cause of kidney disease – insights from Mendelian randomisation studies. medRxiv. 2020:2020.09.13.20155234.

31. Pang Y, Kartsonaki C, Lv J, Millwood IY, Yu C, Guo Y, et al. Observational and Genetic Associations of Body Mass Index and Hepatobiliary Diseases in a Relatively Lean Chinese Population. JAMA Netw Open. 2020;3(10):e2018721.

32. Larsson SC, Burgess S, Michaelsson K. Genetic association between adiposity and gout: a Mendelian randomization study. Rheumatology (Oxford). 2018;57(12):2145–8.

33. Traish AM, Miner MM, Morgentaler A, Zitzmann M. Testosterone deficiency. Am J Med. 2011;124(7):578–87.

34. Basaria S, Coviello AD, Travison TG, Storer TW, Farwell WR, Jette AM, et al. Adverse events associated with testosterone administration. N Engl J Med. 2010;363(2):109–22.

35. Vigen R, O’Donnell CI, Barón AE, Grunwald GK, Maddox TM, Bradley SM, et al. Association of testosterone therapy with mortality, myocardial infarction, and stroke in men with low testosterone levels. Jama. 2013;310(17):1829–36.

36. Ohlander SJ, Varghese B, Pastuszak AW. Erythrocytosis Following Testosterone Therapy. Sex Med Rev. 2018;6(1):77–85.

37. Umer A, Kelley GA, Cottrell LE, Giacobbi P, Jr., Innes KE, Lilly CL. Childhood obesity and adult cardiovascular disease risk factors: a systematic review with meta-analysis. BMC Public Health. 2017;17(1):683.

38. Weihe P, Spielmann J, Kielstein H, Henning-Klusmann J, Weihrauch-Bluher S. Childhood Obesity and Cancer Risk in Adulthood. Curr Obes Rep. 2020;9(3):204–12.

39. Laakso S, Viljakainen H, Lipsanen-Nyman M, Turpeinen U, Ivaska KK, Anand-Ivell R, et al. Testicular Function and Bone in Young Men with Severe Childhood-Onset Obesity. Horm Res Paediatr. 2018;89(6):442–9.

40. Eriksson J, Haring R, Grarup N, Vandenput L, Wallaschofski H, Lorentzen E, et al. Causal relationship between obesity and serum testosterone status in men: A bi-directional mendelian randomization analysis. PLoS One. 2017;12(4):e0176277.

41. Hemani G, Bowden J, Davey Smith G. Evaluating the potential role of pleiotropy in Mendelian randomization studies. Hum Mol Genet. 2018;27(R2):R195-R208.

42. Bowden J, Hemani G, Davey Smith G. Invited Commentary: Detecting Individual and Global Horizontal Pleiotropy in Mendelian Randomization-A Job for the Humble Heterogeneity Statistic? Am J Epidemiol. 2018;187(12):2681–5.

43. Mitchell R, Hemani G, Dudding T, Paternoster L. UK Biobank Genetic Data: MRC-IEU Quality Control, Version 2. data.bris; 2018.

44. Mitchell R, Elsworth B, Mitchell R, Raistrick C, Paternoster L, Hemani G, et al. MRC IEU UK Biobank GWAS pipeline version 2. data.bris; 2019.

