## Supplementary Methods for "Evaluating and implementing block jackknife resampling Mendelian randomization to mitigate bias induced by overlapping samples"

#### UK Biobank study

UK Biobank is a population-based health research resource consisting of approximately 500,000 people, aged between 38 years and 73 years, who were recruited between the years 2006 and 2010 from across the UK (1). Particularly focused on identifying determinants of human diseases in middle-aged and older individuals, participants provided a range of information (such as demographics, health status, lifestyle measures, cognitive testing, personality self-report, and physical and mental health measures) via questionnaires and interviews; anthropometric measures, BP readings and samples of blood, urine and saliva were also taken (data available at www.ukbiobank.ac.uk). A full description of the study design, participants and quality control (QC) methods have been described in detail previously (2). UK Biobank received ethical approval from the Research Ethics Committee (REC reference for UK Biobank is 11/NW/0382).

#### Genotyping and imputation

The full data release contains the cohort of successfully genotyped samples (n=488,377). 49,979 individuals were genotyped using the UK BiLEVE array and 438,398 using the UK Biobank axiom array. Pre-imputation QC, phasing and imputation are described elsewhere (3). In brief, prior to phasing, multiallelic SNPs or those with MAF ≤1 % were removed. Phasing of genotype data was performed using a modified version of the SHAPEIT2 algorithm (4). Genotype imputation to a reference set combining the UK10K haplotype and HRC reference panels (5) was performed using IMPUTE2 algorithms (6). The analyses presented here were restricted to autosomal variants within the HRC site list using a graded filtering with varying imputation quality for different allele frequency ranges. Therefore, rarer genetic variants are required to have a higher imputation INFO score (Info>0.3 for MAF >3%; Info>0.6 for MAF 1-3%; Info>0.8 for MAF 0.5-1%; Info>0.9

for MAF 0.1-0.5%) with MAF and Info scores having been recalculated on an in-house derived 'European' subset (7).

### Data quality control

Individuals with sex-mismatch (derived by comparing genetic sex and reported sex) or individuals with sex-chromosome aneuploidy were excluded from the analysis (n=814). We restricted the sample to individuals of 'European' ancestry as defined by an in-house k-means cluster analysis performed using the first 4 principal components provided by UK Biobank in the statistical software environment R. The current analysis includes the largest cluster from this analysis (n=464,708) (7).

### Association analysis: statistical methods

Genome-wide association analysis (GWAS) was conducted using linear mixed model (LMM) association method as implemented in BOLT-LMM (v2.3) (8). To model population structure in the sample, we used 143,006 directly genotyped SNPs, obtained after filtering on MAF > 0.01; genotyping rate > 0.015; Hardy-Weinberg equilibrium p-value < 0.0001 and LD pruning to an $r^2$ threshold of 0.1 using PLINKv2.00. Genotype array and sex were adjusted for in the model. BOLT-LMM association statistics are on the linear scale. As such, test statistics (betas and their corresponding standard errors) were transformed to log odds ratios and their corresponding 95% confidence intervals on the liability scale using a Taylor transformation expansion series (9).

### References


1. Allen NE, Sudlow C, Peakman T, Collins R, Biobank UK. UK biobank data: come and get it. Sci Transl Med. 2014;6(224):224ed4.
2. Collins R. What makes UK Biobank special? The Lancet. 2012;379(9822):1173-4.
3. Bycroft C, Freeman C, Petkova D, Band G, Elliott LT, Sharp K, et al. The UK Biobank resource with deep phenotyping and genomic data. Nature. 2018;562(7726):203-9.
4. O'Connell J, Sharp K, Shrine N, Wain L, Hall I, Tobin M, et al. Haplotype estimation for biobank-scale data sets. Nat Genet. 2016;48(7):817-20.
5. Huang J, Howie B, McCarthy S, Memari Y, Walter K, Min JL, et al. Improved imputation of low-frequency and rare variants using the UK10K haplotype reference panel. Nat Commun. 2015;6:8111.
6. Howie B, Marchini J, Stephens M. Genotype imputation with thousands of genomes. G3 (Bethesda). 2011;1(6):457-70.
7. Mitchell R, Hemani G, Dudding T, Paternoster L. UK Biobank Genetic Data: MRC-IEU Quality Control, Version 2. data.bris; 2018.
8. Loh PR, Tucker G, Bulik-Sullivan BK, Vilhjalmsson BJ, Finucane HK, Salem RM, et al. Efficient Bayesian mixed-model analysis increases association power in large cohorts. Nat Genet. 2015;47(3):284-90.
9. Loh PR, Kichaev G, Gazal S, Schoech AP, Price AL. Mixed-model association for biobank-scale datasets. Nat Genet. 2018;50(7):906-8.
